# Long-term cognitive recovery after out-of-hospital cardiac arrest: Insights into improvement over six months and the role of arrest duration

**DOI:** 10.1101/2025.03.28.25324863

**Authors:** Jeanet Albøge Enevoldsen Brouer, Lisa Gregersen Oestergaard, Hans Eiskjær, John Bro-Jeppesen, Lola Qvist Kristensen

## Abstract

**Background:** Out-of-hospital cardiac arrest (OHCA) is a significant cause of mortality and morbidity worldwide. While resuscitation advancements have increased survival, many survivors suffer cognitive impairments that affect their quality of life. Most research has focussed on neurological outcomes, while little attention has been paid to cognitive function.

**Aim:** To investigate the proportion of cognitive impairment in OHCA survivors at discharge and six months after cardiac arrest and to investigate the association between the duration of cardiac arrest and level of cognitive function.

**Methods:** In this prospective cohort study, 184 OHCA survivors were assessed using the Montreal Cognitive Assessment (MoCA) screening tool. Duration of cardiac arrest was defined by no-flow, low-flow and time to return of spontaneous circulation (ROSC). Multiple logistic regression analysis provided odds ratios (OR) and confidence intervals (CI).

**Results:** The study indicates a significant improvement in cognitive function among OHCA survivors from discharge to the six-month follow-up. The proportion of patients with normal cognitive function increased from 26% to 67%, while the number of patients with severe and moderate cognitive impairment decreased. These results suggest a general enhancement in cognitive function over time. No significant association was found between the duration of cardiac arrest and cognitive function, either at discharge or follow-up.

**Conclusions:** Cognitive function improved considerably within six months following cardiac arrest, with the proportion of patients exhibiting normal cognitive function increasing from 26% to 67%. This study found no association between the duration of cardiac arrest and cognitive function.

## Introduction

Out-of-hospital cardiac arrest (OHCA) is a major cause of morbidity and mortality worldwide; however, survival rates have improved over the past two decades (1, 2). In Denmark, the 30-day survival rate rose from 4% in 2001 to 14.4% in 2023; worldwide, the 30-day survival ranges from 3% to 18% (3-5). This increase can be attributed to the widespread implementation of prehospital cardiopulmonary resuscitation (CPR) training and increased public access to automated external defibrillators (6), along with advancements in post-resuscitation care following hospital admission (7, 8).

Hypoxic brain injury is a common sequelae of cardiac arrest (9-12), often leading to cognitive impairments ranging from mild to severe (13-15). These impairments occur primarily in the domains of memory, attention and/or executive function, depending on the region of injury (16, 17). Most resuscitation science has focused on survival and survival with favourable neurological outcomes, typically assessed using the five-point Cerebral Performance Category (CPC) scale or the six-point modified Rankin Scale (mRS). However, knowledge is limited about the specific cognitive impairments commonly observed among OHCA survivors, which may have significant implications on daily activities. Previous studies have explored the association between the duration of cardiac arrest and the neurological outcomes in OHCA survivors; however, they have not specifically focused on cognitive function (18-21).

The duration of the cardiac arrest is widely recognised as an important factor influencing survival and neurological outcomes following OHCA. A cohort study (N=11,368) demonstrated a steady decline in the likelihood of survival with a favourable neurological outcome (mRS ≤3) at hospital discharge as the duration of professional CPR increased (18).

The duration of cardiac arrest can be defined by two distinct intervals: 1) no-flow (interval from collapse to initiation of CPR) and 2) low-flow (interval from start of CPR to return of spontaneous circulation (ROSC) or termination of resuscitation (19, 22). Despite a strong association between the duration of OHCA and an adverse neurological outcome, complete recovery after prolonged CPR may be achieved in selected cases (23, 24).

Research indicates that approximately 50% of OHCA survivors achieve a CPC score of <2, which is internationally recognised as a successful outcome. However, almost half of these survivors later experience cognitive impairments, highlighting the complexity and challenges in accurately evaluating long-term outcomes (26, 27).

Limited knowledge exists regarding the relationship between the duration of cardiac arrest and cognitive function, especially when assessed using the Montreal Cognitive Assessment (MoCA) screening tool.

This study aimed to investigate the proportion of cognitive impairment at hospital discharge and its progression over six months following cardiac arrest. Additionally, it sought to investigate the association between the duration of cardiac arrest and level of cognitive function at both hospital discharge and six-month follow-up in a population of OHCA survivors. We hypothesised that a longer duration of cardiac arrest would be associated with poorer cognitive outcomes.

## Materials and methods

### Design, setting and participants

This study was a prospective single-centre observational cohort study with a follow-up period of six months after cardiac arrest. The study was conducted as a sub-study of an ongoing cohort study investigating activities of daily living after surviving cardiac arrest (25). The present study utilises data on the duration of cardiac arrest and cognitive function collected from this more extensive study. Patients were recruited from the Department of Cardiology at Aarhus University Hospital, Denmark, between June 2019 and March 2023, and their data were collected after verbal and written consent had been obtained. Inclusion criteria were successful resuscitation from OHCA, age ≥18 years and sufficient command of the Danish language to understand participant information and to give informed consent, based on the occupational therapist’s assessment. Participants were excluded if no data were available on the duration of cardiac arrest or the assessment of cognitive function (Fig. 1). The study was registered at the Central Denmark Region (1-16-02-261-19). Additionally, the Ethics Committee waived the need for further registration of the study (1-10-72-1-20). Furthermore, this study was conducted in accordance with the Strengthening the Reporting of Observational Studies in Epidemiology (STROBE) statement (26).

**Fig. 1:**
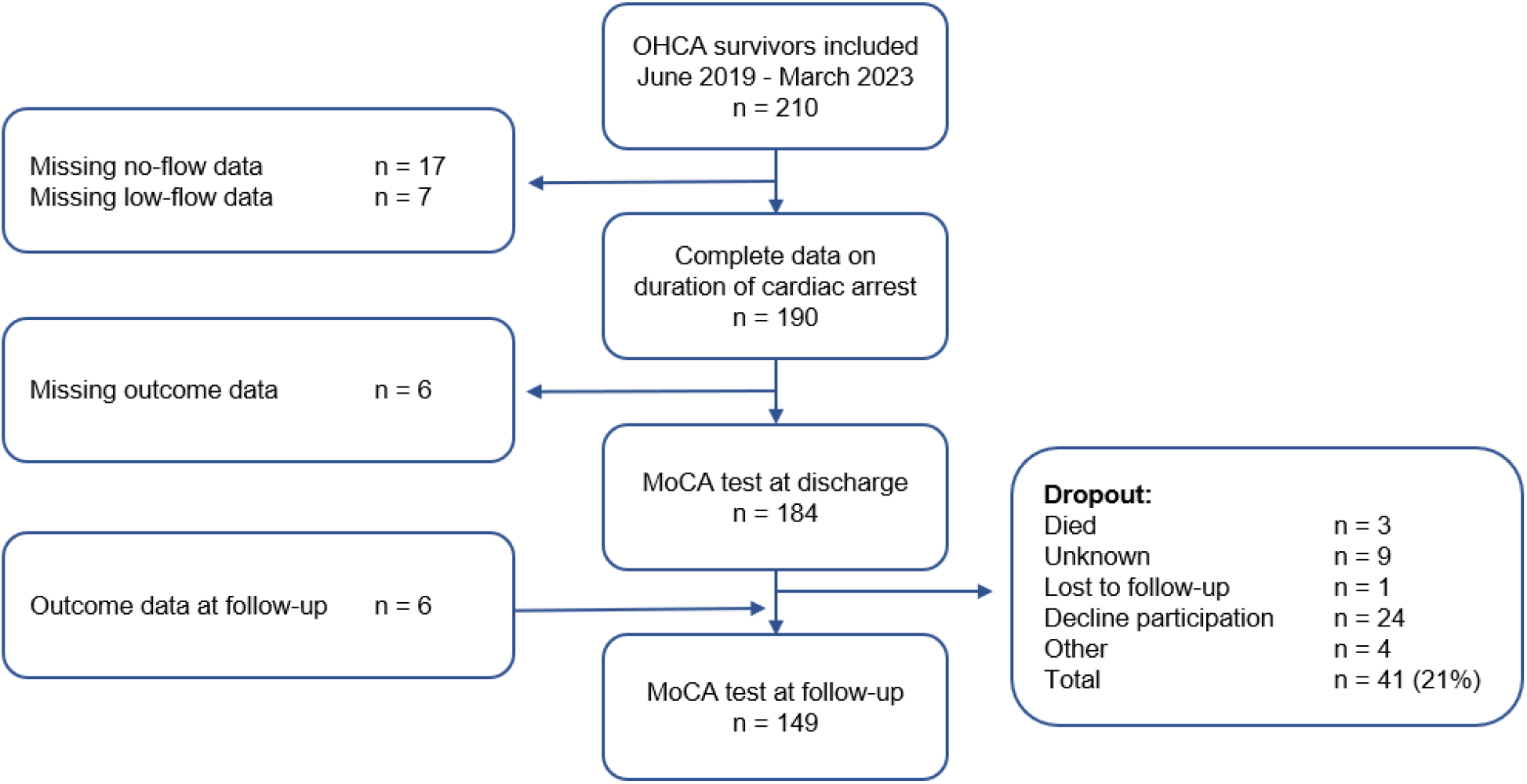
Flowchart of participation in the studys Abbreviations. OHCA: Out-of-hospital cardiac arrest. MoCA: Montreal Cognitive Assessment.

### Data collection

#### Demographics and clinical variables

Sociodemographic data, such as marital status, education and employment status, were self-reported. Data on age, sex, comorbidities, witnessed arrest, bystander CPR, primary rhythm and location were extracted from the Electronic Patient Journal and the Central Prehospital Patient Journal from Aarhus University Hospital, Denmark, at baseline.

#### Duration of cardiac arrest

The primary independent variable of interest was the duration of cardiac arrest measured by no-flow and low-flow. No-flow was defined as the time interval in minutes from the bystander emergency call (recorded when the call was answered at the emergency services and assumed to be the approximate arrest time) to the CPR initiation. Low-flow duration was defined as the time in minutes from the start of CPR to ROSC. The total duration until ROSC was achieved was calculated by adding no-flow and low-flow times. Data were collected from the Electronic Patient Journal and the Prehospital Patient Journal, which were reported by the prehospital emergency physician.

#### The Montreal Cognitive Assessment (MoCA)

Cognitive function was assessed within the last 5 days before discharge from the hospital and repeated at approximately six months (+/-14 days) after cardiac arrest, both assessed by the MoCA tool.

The MoCA is a widely used screening tool for mild cognitive impairment. It assesses seven cognitive domains: Visuospatial and executive abilities, naming, attention, language, abstraction, delayed recall and orientation (27). Each cognitive domain is assigned a specific score, which is summed to produce a total score ranging from 0 to 30 points. The test can be administered in approximately 10-15 minutes and is a brief paper-and-pencil cognitive screening tool. The MoCA score is adjusted for educational level by awarding one extra point to anyone with 12 or fewer years of formal education. This adjustment aims to minimise the risk of classifying less educated subjects as cognitively impaired. A cut-off of ≥26 is considered normal cognitive function (27). Both baseline and follow-up assessments were conducted by one of four certified and experienced occupational therapists at the Department of Cardiology, Aarhus University Hospital.

### Statistical methods

Descriptive statistics were used to present participants’ baseline characteristics. The distribution of data was presented by frequencies (n) and proportions (%) for categorical variables and by means and standard deviations (SD) for continuous variables if normally distributed or otherwise as medians with quartiles (Q).

The analysis was performed by logistic regression, calculating odds ratio (OR) and 95% confidence intervals (CI) to estimate the odds of a cognitive function below the cut-off (<26) on MoCA after cardiac arrest. Secondarily, the association between the duration of cardiac arrest and cognitive function after six months was examined.

Both crude and adjusted ORs were calculated for the associations. We adjusted for potential confounders (if no interactions were identified) such as age, sex, comorbidities, witnessed arrest, bystander CPR, the initial recorded rhythm and the location of the arrest. A p-value of <0.05 was considered statistically significant. All analyses and data management were conducted in STATA version 18.0 (StataCorp, College Station, Texas, USA).

## Results

### Study population

Among admitted survivors after OHCA, who were alive at hospital discharge, 210 met the inclusion criteria. As shown in the flowchart (Fig. 1), data on the duration of cardiac arrest were available for a study population of 190 participants. Of those, MoCA scores were available for 184 participants. During the six months before follow-up, another 21% dropped out for various reasons. Data regarding MoCA were available at follow-up in 149 participants (Fig. 1).

Overall patient characteristics are shown in Table 1. The participants’ mean age was 63 years (SD ± 13), with 83% being male. The proportion of witnessed arrests was 96%, and 91% received bystander CPR. Ventricular fibrillation/ventricular tachycardia (VF/VT) was the initial rhythm in 86% of the participants. Regarding marital status, 74% were living with a spouse. Retirees accounted for 47%, and 41% were employed full-time before their cardiac arrest. Cardiovascular comorbidities were present in 44%. Notably, 88% of the participants were referred for rehabilitation upon discharge.

**Table 1:**
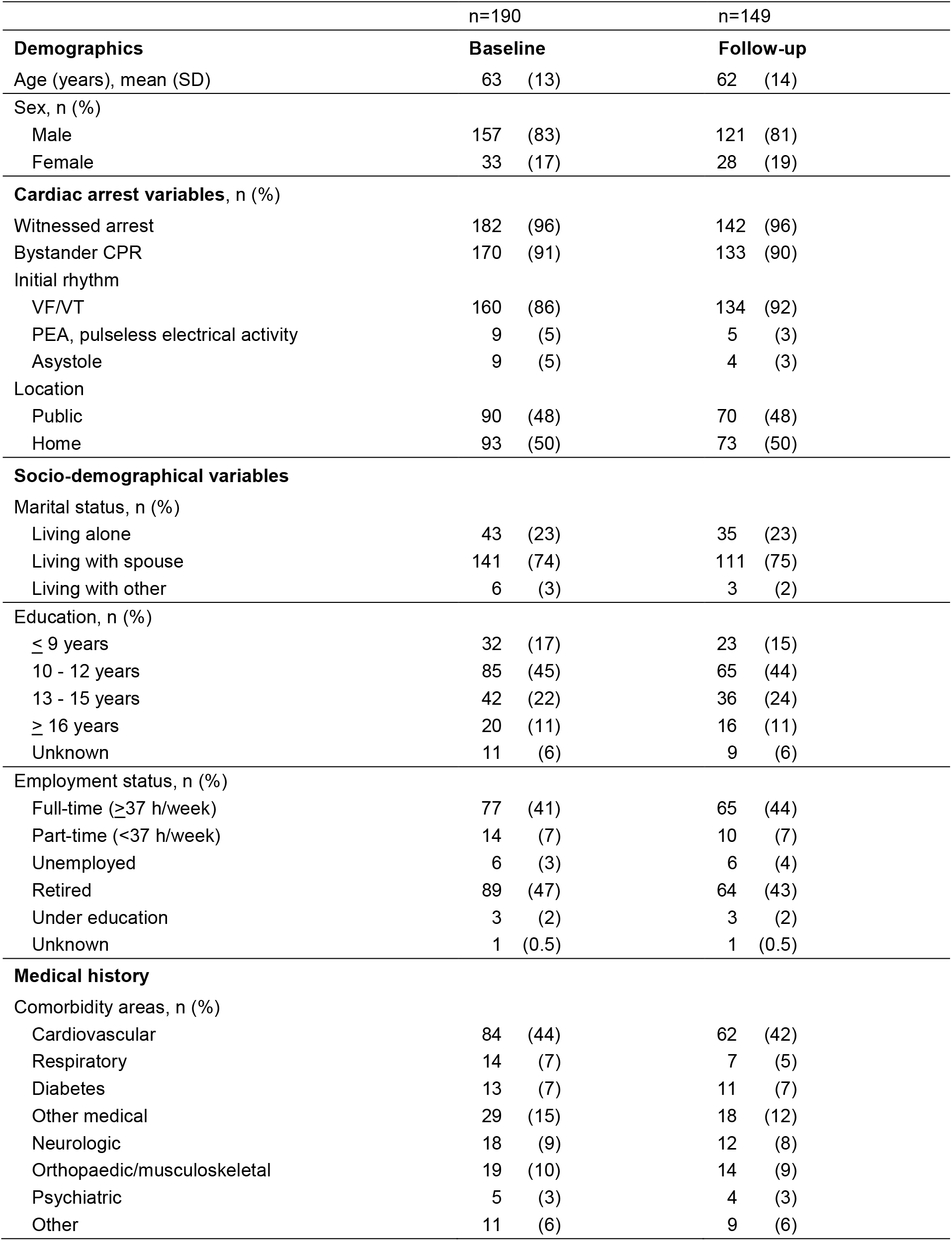
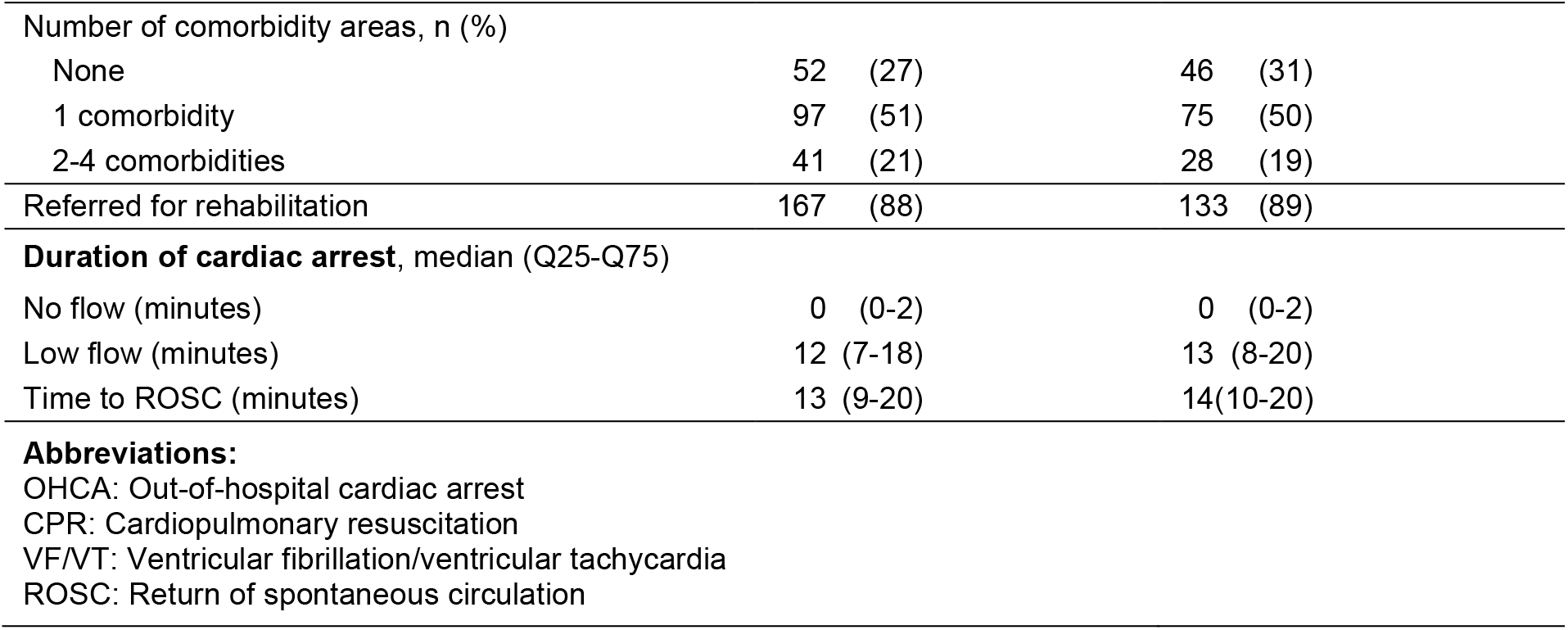
Characteristics of OHCA survivors at discharge and six-month follow-up.

|  | n=190 |  | n=149 |  |
| --- | --- | --- | --- | --- |
| **Demographics** | **Baseline** |  | **Follow-up** |  |
| Age (years), mean (SD) | 63 | (13) | 62 | (14) |
| Sex, n (%) |  |  |  |  |
| Male | 157 | (83) | 121 | (81) |
| Female | 33 | (17) | 28 | (19) |
| **Cardiac arrest variables, n (%)** |  |  |  |  |
| Witnessed arrest | 182 | (96) | 142 | (96) |
| Bystander CPR | 170 | (91) | 133 | (90) |
| Initial rhythm |  |  |  |  |
| VF/VT | 160 | (86) | 134 | (92) |
| PEA, pulseless electrical activity | 9 | (5) | 5 | (3) |
| Asystole | 9 | (5) | 4 | (3) |
| Location |  |  |  |  |
| Public | 90 | (48) | 70 | (48) |
| Home | 93 | (50) | 73 | (50) |
| **Socio-demographical variables** |  |  |  |  |
| Marital status, n (%) |  |  |  |  |
| Living alone | 43 | (23) | 35 | (23) |
| Living with spouse | 141 | (74) | 111 | (75) |
| Living with other | 6 | (3) | 3 | (2) |
| Education, n (%) |  |  |  |  |
| ≤ 9 years | 32 | (17) | 23 | (15) |
| 10 - 12 years | 85 | (45) | 65 | (44) |
| 13 - 15 years | 42 | (22) | 36 | (24) |
| ≥ 16 years | 20 | (11) | 16 | (11) |
| Unknown | 11 | (6) | 9 | (6) |
| Employment status, n (%) |  |  |  |  |
| Full-time (≥37 h/week) | 77 | (41) | 65 | (44) |
| Part-time (<37 h/week) | 14 | (7) | 10 | (7) |
| Unemployed | 6 | (3) | 6 | (4) |
| Retired | 89 | (47) | 64 | (43) |
| Under education | 3 | (2) | 3 | (2) |
| Unknown | 1 | (0.5) | 1 | (0.5) |
| **Medical history** |  |  |  |  |
| Comorbidity areas, n (%) |  |  |  |  |
| Cardiovascular | 84 | (44) | 62 | (42) |
| Respiratory | 14 | (7) | 7 | (5) |
| Diabetes | 13 | (7) | 11 | (7) |
| Other medical | 29 | (15) | 18 | (12) |
| Neurologic | 18 | (9) | 12 | (8) |
| Orthopaedic/musculoskeletal | 19 | (10) | 14 | (9) |
| Psychiatric | 5 | (3) | 4 | (3) |
| Other | 11 | (6) | 9 | (6) |

|  |  |  |  |
| --- | --- | --- | --- |
| Number of comorbidity areas, n (%) |  |  |  |
| None | 52 | (27) | 46 (31) |
| 1 comorbidity | 97 | (51) | 75 (50) |
| 2-4 comorbidities | 41 | (21) | 28 (19) |
| Referred for rehabilitation | 167 | (88) | 133 (89) |
| **Duration of cardiac arrest, median (Q25-Q75)** |  |  |  |
| No flow (minutes) | 0 | (0-2) | 0 (0-2) |
| Low flow (minutes) | 12 | (7-18) | 13 (8-20) |
| Time to ROSC (minutes) | 13 | (9-20) | 14(10-20) |
| **Abbreviations:** |  |  |  |
| OHCA: Out-of-hospital cardiac arrest |  |  |  |
| CPR: Cardiopulmonary resuscitation |  |  |  |
| VF/VT: Ventricular fibrillation/ventricular tachycardia |  |  |  |
| ROSC: Return of spontaneous circulation |  |  |  |

Data on no-flow and low-flow time were available for 190 participants. The median of no-flow time was 0 minutes (0-2) with a range of 0 to10 minutes; the median observed low-flow was 12 minutes (7-18) with a range of 1 to 102 minutes; and the median of time to ROSC was 13 minutes (9-20).

### Cognitive function at discharge and development over six months

Table 2 shows the cognitive function at discharge and temporal changes over the course of the next six months after cardiac arrest in OHCA survivors. A total of 26% had normal cognitive function at hospital discharge, indicated by a MoCA ≥26 score. The median total MoCA score at discharge was 23 (20-26), with scores ranging from 8 to 30 points. Most patients (59%) had a mildly reduced MoCA score (Table 2).

**Table 2.** Prevalence of cognitive impairment at discharge and six-month follow-up, measured by MoCA and divided into four groups.

|  | n=184 |  | n=149 |  |
| --- | --- | --- | --- | --- |
| **Cognitive function** | **Baseline** |  | **Follow-up** |  |
| Severe 0-9, n (%) | 2 | (1) | 0 | (0) |
| Moderate 10-17, n (%) | 25 | (14) | 6 | (4) |
| Mild 18-26, n (%) | 108 | (59) | 44 | (29) |
| Normal 26-30, n (%) | 49 | (26) | 99 | (67) |
| Total MoCA score, median (Q25-Q75) | 23 | (20-26) | 26 | (24-28) |
| **Abbreviations:** |  |  |  |  |
| MoCA: Montreal Cognitive Assessment |  |  |  |  |

At the six-month follow-up, a dropout rate of 21% was observed, with the majority citing lack of energy as the primary reason for non-participation (Fig. 1). Most of those who dropped out had a MoCA score below 26 at discharge.

At the six-month follow-up, 67% of the participants had normal cognitive function. The median MoCA score at follow-up was 26 (24-28), ranging from 12 to 30 points (Table 2). Only 4% of the patients had a moderately reduced and none a severely reduced MoCA score.

Figure 2 presents a Sankey diagram illustrating the distribution of MoCA scores across four severity levels at baseline and six months after cardiac arrest, illustrating changes in cognitive function over time. Complete MoCA data at both discharge and the six-month follow-up were available for 143 participants.

**Fig. 2.**
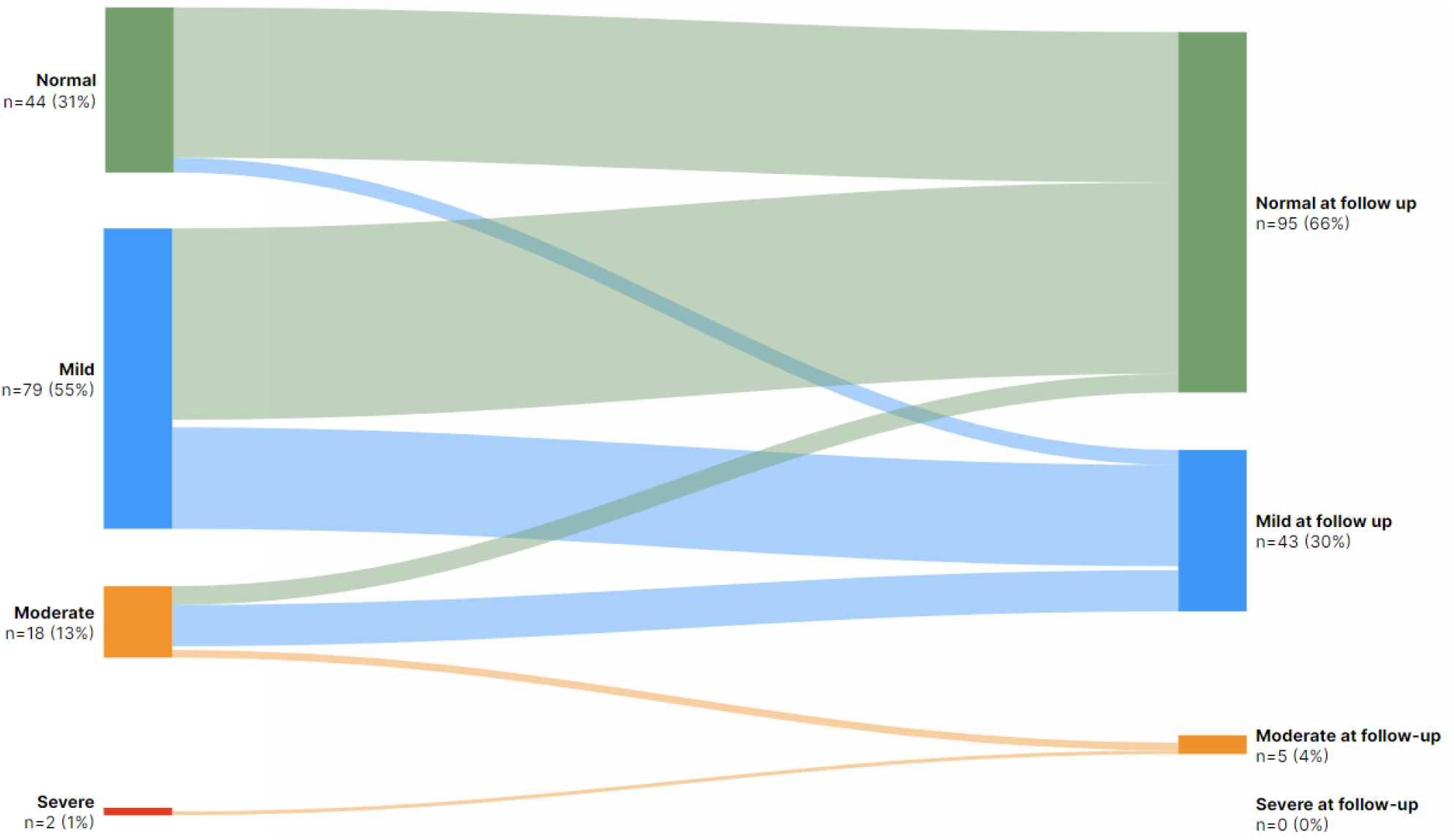
Changes in MoCA between discharge and follow-up six months after cardiac arrest for those where both measurements were present (n = 143). MoCA score is divided into four groups at baseline and six months after cardiac arrest: Green: 26-30 = normal cognitive function Blue: 18-25 = mild cognitive impairment Orange: 10-17 = moderate cognitive impairment Red: 0-9 = severe cognitive impairment

After six months, the proportion of OHCA survivors with normal cognitive function (MoCA ≥26) increased significantly, from 31% at discharge to 67%. Approximately one-third of survivors had mild cognitive impairment at the six-month follow-up. The majority of patients (n=18/20, 90%) with moderate or severe cognitive impairment at discharge demonstrated improved cognitive status at six months.

Figure 3 reveals the seven cognitive MoCA domains, each with its percentage of the maximum possible score in each domain. An increase in scores is seen at six months of follow-up. Among the cognitive domains assessed, memory was the lowest-scoring domain for participants, while naming, attention and orientation had the highest scores.

**Fig. 3.**
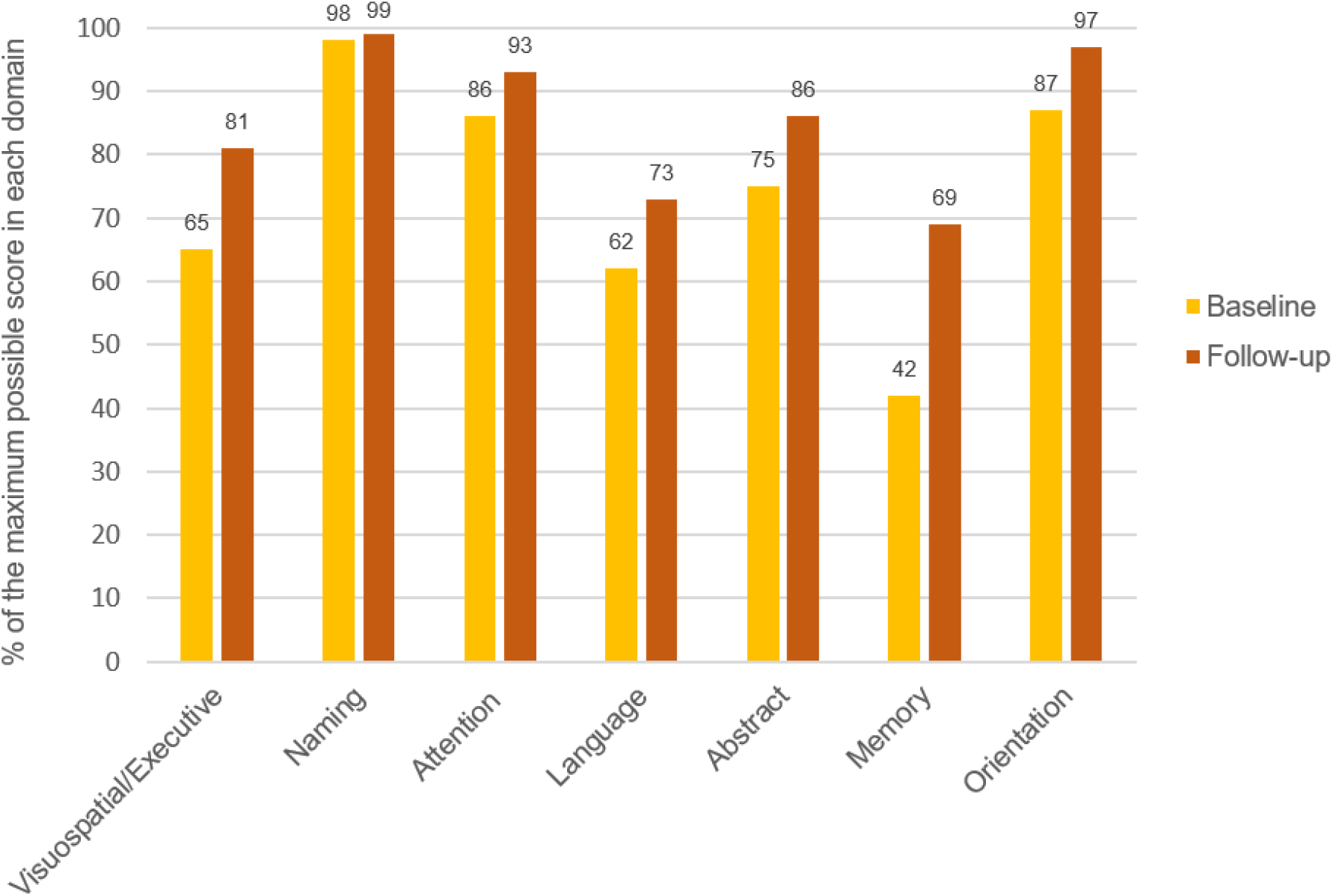
MoCA score among different cognitive domains at hospital discharge and six months after cardiac arrest

### Association between duration of cardiac arrest and cognitive function at hospital discharge

We found no association between the duration of cardiac arrest and cognitive function at hospital discharge. The association between the duration of cardiac arrest and cognitive function did not change when adjusting for age, sex, comorbidities, witnessed arrest, bystander CPR, the initial recorded rhythm and location of arrest, Table 3.

**Table 3:** Logistic regression of the association between no-flow and low-flow (duration of cardiac arrest) and cognitive function measured with MoCA<26 at hospital discharge.

| n = 184 | Crude |  |  | Adjusted |  |  |
| --- | --- | --- | --- | --- | --- | --- |
|  | OR | 95% CI | p-value | OR | 95% CI | p-value |
| No-flow | 1.12 | 0.91; 1.37 | 0.286 | 0.83 | 0.61; 1.13 | 0.241 |
| Low-flow | 0.99 | 0.96; 1.02 | 0.561 | 0.99 | 0.96; 1.03 | 0.739 |
| Time to ROSC | 0.99 | 0.97; 1.02 | 0.683 | 0.99 | 0.96; 1.03 | 0.654 |

Adjusted for age, sex, comorbidities (yes/no), witnessed arrest, bystander CPR, the initial recorded rhythm and location of cardiac arrest.

The association between no-flow and cognitive function at hospital discharge showed an unadjusted OR of 1.12 (95% CI: 0.91; 1.37) and an adjusted OR of 0.83 (95% CI: 0.61; 1.13). Similarly, the association between low-flow duration and cognitive function revealed an unadjusted OR of 0.99 (95% CI: 0.96; 1.02) and an adjusted OR of 0.99 (95% CI: 0.96; 1.03). The association between the total duration until ROSC and cognitive function showed an unadjusted OR of 0.99 (95% CI: 0,97; 1,02) and an adjusted OR of 0.99 (95% CI: 0.96; 1.03). None of the results were statistically significant (Table 3).

### Association between duration of cardiac arrest and cognitive function after six months

Table 4 presents the results of the analysis, revealing that among OHCA survivors, no association was observed between the duration of cardiac arrest and subsequent cognitive function at six months after cardiac arrest. Adjusting for age, sex, comorbidities, witnessed arrest, bystander CPR, the initial recorded rhythm and location of arrest did not change this result.

**Table 4:**
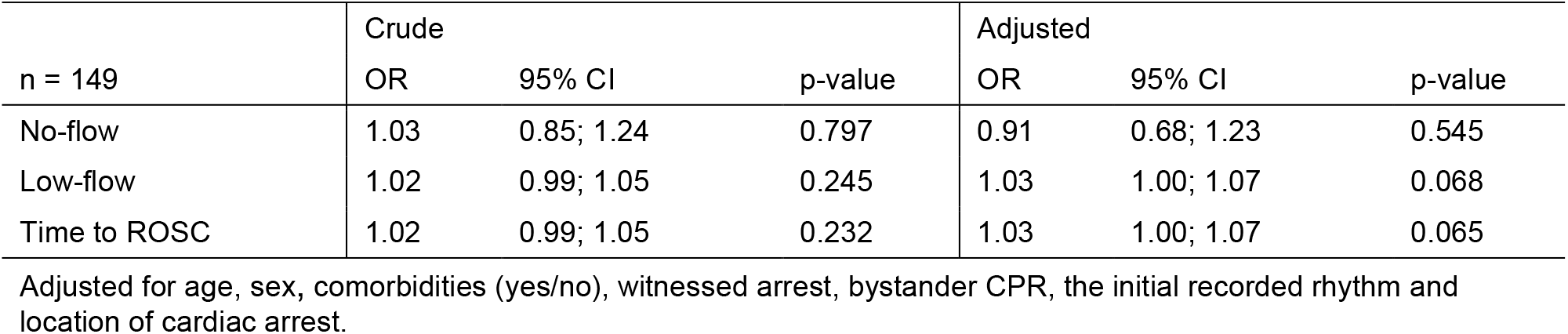
Logistic regression of the association between no-flow and low-flow (duration of cardiac arrest) and cognitive function measured with MoCA<26 six months after cardiac arrest.

The association between no-flow and cognitive function at discharge showed an unadjusted OR of 1.03 (95% CI: 0.85; 1.24) and an adjusted OR of 0.91 (95% CI: 0.68; 1.23). For low-flow duration, the unadjusted OR was 1.02 (95% CI: 0.99; 1.05), and the adjusted OR was 1.03 (95% CI: 1.00; 1.07). Similarly, the association between total duration until ROSC was achieved and cognitive function had an unadjusted OR of 1.02 (95% CI: 0.99; 1.05) and an adjusted OR of 1.03 (95% CI: 1.00; 1.07). None of the results were statistically significant.

## Discussion

This study investigated 184 adult OHCA survivors, revealing that 26% had normal cognitive function at hospital discharge. Six months after cardiac arrest, 67% of the 149 participants showed normal cognitive function (MoCA > 26). Concurrently, the prevalence of severe and moderate cognitive impairments decreased. These findings indicate a significant improvement in cognitive function among OHCA survivors from discharge to the six-month follow-up.

Contrary to our study hypothesis, our findings revealed no statistically significant association between the duration of cardiac arrest and the level of cognitive function at either hospital discharge or six months after cardiac arrest in patients surviving hospital discharge. These findings suggest that the duration of cardiac arrest does not necessarily affect cognitive function, either at hospital discharge or after six months, in a population of patients with witnessed OHCA who received almost immediate CPR.

To our knowledge, this is the first study to investigate the association between the duration of cardiac arrest and cognitive function using the MoCA. Previous research has focused on survival and neurological outcomes, measured with CPC or mRS, rather than cognitive function specifically (12, 28). The high proportion of OHCA survivors who subsequently experienced cognitive impairment emphasises the importance of assessing cognitive function separately from overall neurological function in OHCA survivors. The present study aimed to address this gap in the literature by examining cognitive function using MoCA.

The findings of this study differ in certain respects from previous neurologically focused research. Reynolds et al. reported a decline in favourable neurological outcomes at hospital discharge with increasing CPR duration; however, they also noted a relatively high incidence of complete recovery after prolonged CPR (18). In contrast, Mion et al. found no association between the duration of cardiac arrest and cognitive impairment at discharge (29).

Differences in study populations and definitions of cardiac arrest duration may explain some of these discrepancies. For instance, Guy et al.’s study of over 7,000 OHCA cases demonstrated that no-flow duration significantly affects the likelihood of favourable OHCA outcomes at the time of hospital discharge in North America (20). They found that for each additional minute of no-flow, the adjusted risk of a favourable neurologic outcome decreased by 13%. Notably, no patients with a no-flow duration exceeding 20 min achieved a favourable neurological outcome. To ensure the validity of no-flow estimates, they excluded patients if the arrest was unwitnessed or if CPR or a defibrillator was administered by bystanders. In contrast, our study did not exclude unwitnessed arrests or differentiate between bystander and emergency medical personnel-administered CPR. Additionally, participants were included only after being transferred to the cardiology ward, regardless of their prior stay in the intensive care unit. This may have led to a selected population in our study. This difference in reported results may be attributed to differences in study populations along with variations in sample sizes between the studies.

Assessing the duration of cardiac arrest in OHCA can be challenging and is often subject to some degree of imprecision, as different time estimation methods are used across studies, which may affect the reported accuracy of the reported durations (30). Additionally, witnesses may vary in the accuracy and reliability of their recollections of event timings, which can further influence the reported no-flow and low-flow durations (31). In addition, low-flow time is generally assumed to represent high-quality compressions with minimal interruptions, even when CPR is performed by a bystander. Furthermore, differences in how patients tolerate low-flow conditions are influenced by factors such as age and pre-existing health conditions, which affect the comparability of data between studies (22).

### Clinical impact

The findings of this study have important implications for clinical practice. Our results demonstrate that prolonged cardiac arrest in survivors does not necessarily correlate with impaired cognitive function, either in the short or long term. This challenges previous assumptions regarding the negative impact of extended resuscitation efforts on the risk of severe cognitive impairment. This suggests that clinicians should avoid making premature judgments about the cognitive prognosis for patients who undergo prolonged CPR if cardiac arrest has been witnessed and CPR started promptly. Consequently, optimism about potential cognitive recovery can be maintained, and efforts to optimise patient care should focus on early identification and intervention for cognitive impairments, irrespective of arrest duration.

Routine cognitive screening of OHCA survivors using MoCA is recommended by the European Resuscitation Council and the European Society of Intensive Care Medicine to detect early signs of cognitive impairment (11). MoCA has proven to be a valuable tool in this context, with a cutoff score of 26 providing a reliable indicator of cognitive impairment (27). The test has demonstrated high sensitivity (90%) and specificity (87%) for identifying mild cognitive impairment, offering clinicians confidence in its utility (27, 32, 33). Moreover, research has shown that MoCA has a strong construct validity (34) and good internal consistency (Cronbach’s alpha = 0.83), indicating its reliability (27) and accuracy in measuring cognitive impairment.

Screening should ideally occur both before discharge and at follow-up to observe long-term cognitive recovery (11). Early identification of cognitive impairment is essential as it allows for referral to a targeted cognitive rehabilitation programme. Our results suggest cognitive rehabilitation programmes should prioritise memory training, concentration exercises and strategies to reduce cognitive fatigue, as these areas are most often impacted.

As survival rates improve, cognitive outcomes are becoming an increasingly critical component of post-resuscitation care and a key factor in restoring daily life activities. Early identification of cognitive impairments may support clinicians in tailoring rehabilitation interventions to address specific deficits, with the potential to improve everyday quality of life after discharge. The use of MoCA provides a standardised and validated approach to cognitive assessment, supported by evidence of its sensitivity, specificity and predictive values. However, it does not measure processing speed, which is a limitation.

### Strengths and limitations

A key strength of our study is its prospective design, allowing for cognitive assessment at two time points: hospital discharge and six months after cardiac arrest. This longitudinal approach captures both immediate and longer-term cognitive outcomes. Furthermore, MoCA provides a detailed evaluation across cognitive domains, enhancing our understanding of specific impairments in OHCA survivors (7, 27).

In Denmark, prehospital care systematically documents no-flow and low-flow times, including cases where resuscitation is initiated by witnesses. All data are validated in the prehospital database to ensure high data quality. Additionally, Denmark places a strong emphasis on promoting bystander intervention during cardiac arrests, as early bystander CPR can significantly improve survival rates. CPR training has been provided in schools and workplaces, and many individuals have registered as volunteer first responders (“heart runners”). This effort has led to a high percentage of cases where CPR is initiated immediately by witnesses, resulting in minimal no-flow time. In our study population, 61% of cases had zero minutes of no-flow, indicating that CPR was initiated immediately.

However, our study also has limitations. Although we adjusted for potential confounders, such as age, sex, comorbidities, and witnessed arrest, residual confounding might still have influenced the results. Factors such as inpatient interventions or other unmeasured variables could also play a role. As mentioned, measuring time intervals during OHCA may be challenging, and any time measurement from bystander CPR until the arrival of the emergency medical services may be inaccurate.

A dropout rate of 21% at the six-month follow-up, with the majority citing a lack of energy as the primary reason for non-participation (Fig. 1), may have impacted the results. Of note, most individuals in the dropout group had a MoCA score below 26 at discharge, which could potentially skew the observed improvement in cognitive function at follow-up.

The relationship between the duration of cardiac arrest and cognitive function is complex, and further research is required to better understand which factors influence cognitive impairments in OHCA survivors in order to devise targeted interventions aimed at improving long-term cognitive function and quality of life for this population. The results from this study may support clinicians managing cardiac arrest patients with the knowledge that cognitive function is not necessarily adversely affected by prolonged no-flow and low-flow durations and that temporal improvements can be achieved within six months.

## Conclusion

This study investigated the proportion of cognitive impairment at discharge and its progression over six months following cardiac arrest. Additionally, we explored the association between the duration of cardiac arrest and cognitive function at both hospital discharge and six-month follow-up in OHCA survivors.

While only 26% of survivors had normal cognitive function at discharge, 67% achieved normal cognitive function after six months. Moreover, an improvement was observed among those who scored moderate cognitive function (MoCA score 11-17). These results indicate that cognitive recovery may occur in the majority of this selected group of OHCA survivors, regardless of the duration of cardiac arrest. Contrary to our hypothesis, we found no significant association between the duration of cardiac arrest and cognitive function assessed by MoCA.

## Data Availability

Data is available in an anonymized form

